# Identification of Novel Syncytiotrophoblast Membrane Extracellular Vesicles Derived Protein Biomarkers in Early-onset Preeclampsia: A Cross-Sectional Study

**DOI:** 10.1101/2023.06.03.23290935

**Authors:** Toluwalase Awoyemi, Shuhan Jiang, Bríet Bjarkadóttir, Maryam Rahbar, Prasanna Logenthiran, Gavin Collett, Wei Zhang, Adam Cribbs, Ana Sofia Cerdeira, Manu Vatish

## Abstract

**Background:** Preeclampsia (PE), a multi-systemic hypertensive pregnancy disease that affects 2-8% of pregnancies worldwide, is a leading cause of adverse maternal and fetal outcomes. Current clinical PE tests have a low positive predictive value for PE prediction and diagnosis. The placenta notably releases extracellular vesicles from the syncytiotrophoblast (STB-EV) into the maternal circulation.

**Objective:** To identify a difference in placenta and STB-EV proteome between PE and normal pregnancy (NP), which could lead to identifying potential biomarkers and mechanistic insights.

**Methods:** Using ex-vivo dual lobe perfusion, we performed mass spectrometry on placental tissue, medium/large and small STB-EVs isolated from PE (n = 6) and NP (n = 6) placentae. Bioinformatically, mass spectrometry was used to identify differentially carried proteins. Western blot was used to validate the identified biomarkers. We finished our investigation with an in-silico prediction of STB-EV mechanistic pathways.

**Results:** We identified a difference in the STB-EVs proteome between PE and NP. Filamin B, collagen 17A1, pappalysin-A2, and scavenger Receptor Class B Type 1) were discovered and verified to have different abundances in PE compared to NP. In silico mechanistic prediction revealed novel mechanistic processes (such as abnormal protein metabolism) that may contribute to the clinical and pathological manifestations of PE.

**Conclusions:** We identified potentially mechanistic pathways and identified differentially carried proteins that may be important in the pathophysiology of PE and are worth investigating because they could be used in future studies of disease mechanisms and as biomarkers.

**Funding:** This research was funded by the Medical Research Council (MRC Programme Grant (MR/J0033601) and the Medical & Life Sciences translational fund (BRR00142 HE01.01)

## Introduction

Preeclampsia (PE) is a significant cause of maternal and neonatal morbidity and mortality, affecting 2-8% of all pregnancies(Lisonkova & Joseph, 2013). It is characterized by hypertension (systolic blood pressure ≥ 140mmHg / diastolic pressure ≥ 90mmHg), and either proteinuria (protein/creatinine ratio of ≥ 30 mg/mmol or more), or evidence of maternal acute kidney injury, liver dysfunction, neurological abnormalities, hemolysis, or thrombocytopenia, and/or fetal growth restriction.(“ACOG Practice Bulletin No. 202: Gestational Hypertension and Preeclampsia,” 2019; Brown et al., 2018) Predicting or early detection of PE is thus of extreme importance to reduce the chance of long term complications but this has been challenging due to the limitations of current predictive models and biochemical tests (which lack in positive predictive value)(Zeisler et al., 2016). The existing tests perform far better in ruling out rather than ruling in PE. They are also most effective shortly before the onset of the disease and within a specific time frame (1 or 2 weeks) rather than earlier in pregnancy(Thadhani et al., 2022).

The pathophysiology of PE implicates the placenta. It is known that PE can occur in trophoblastic tumors (without the presence of a fetus); that PE is more common in multiple pregnancy (with greater placental mass) and that it has occurred in ectopic pregnancies (excluding the involvement of the uterus)(Billieux et al., 2004; Hailu et al., 2017; C. W. G. Redman et al., 2022; Soto-Wright et al., 1995). Finally, delivery of the placenta (irrespective of gestational age) is currently the only cure for the condition(C. Redman, 2014).

The placental syncytiotrophoblast (STB) layer, the interface between the fetus and the mother which lies in direct contact with the maternal circulation. EVs, including STB-EVs, are membrane-bound and cell-derived particles that carry different cargos, including proteins, ribonucleic acid (RNA), deoxyribonucleic acid (DNA) and lipids(Raposo & Stoorvogel, 2013). EVs are based on size into medium/large STB-EVs (MVs, 201–1000nm) or small STB-EVs (≤200nm)(Dragovic et al., 2015a). The release of syncytiotrophoblast extracellular vesicles (STB-EVs), including both size ranges, into the maternal circulation increases with gestation and is further elevated in PE (Germain et al., 2007; Goswamia et al., 2006).Variations in cargo content are evident among different subsets of syncytiotrophoblast-derived extracellular vesicles (STB-EVs), with medium/large STB-EVs (m/lSTB-EVs) showing higher levels of total RNA and total protein in comparison to small STB-EVs (sSTB-EVs)(Zabel et al., 2021). Additionally, distinct miRNA profiles have been noted to differ in abundance between these subsets. While limited studies have investigated disparities in EV subtypes, Keerthikumar et al reported that exosomes and other small extracellular vesicles (EVs) have a more pronounced impact on cell migration and proliferation than the larger EVs known as ectosomes (Keerthikumar et al., 2015) . Furthermore, Minciacchi et al demonstrated that large EVs exhibit greater efficiency in reprogramming fibroblasts and promoting the formation of endothelial cell tubes when compared to small EVs(Minciacchi et al., 2017).

In addition, when it comes to immune cell-derived EVs, large EVs were found to induce the secretion of Th2-associated cytokines, while both medium and small EVs (pelleted at 10,000g and 100,000g) triggered the release of Th1 cytokines(Tkach et al., 2017).These functional distinctions were also observed in STB-EVs. Specifically, our investigation unveiled that normal pregnancy (NP) medium/large-sized STB-EVs significantly enhanced the transcriptional expression of pro-inflammatory cytokines in contrast to pathological (PE) medium/large-sized STB-EVs(Awoyemi et al., 2021). However, there was no significant difference in the small STB-EV population between the two groups, although, in general, NP small STB-EVs exhibited a slight upregulation of the same cytokines(Awoyemi et al., 2021). It is reasonable to conduct further in-depth exploration into the qualitative functional properties of distinct STB-EVs, with the initial step being a comprehensive characterization of these diverse subtypes. The presence of proteins, RNA species and DNA in EVs combined with their constitutional release as inter-cellular signaling moieties means they also have potential in disease prediction and diagnosis. Notably, these distinct STB-EV subtypes have also been identified in the bloodstream. Nakahara et al extensive review compiles a collection of studies that have specifically isolated various subtypes of EVs from maternal plasma(Nakahara et al., 2020). In this study, we performed proteomic analysis of placenta and STB-EVs in PE and NP to identify potential placenta-derived biomarkers. We also conducted in silico analysis on the proteomes to detect potential molecular targets, mechanisms, and processes of PE.

## Methods

### Ethics approval and patient information

Oxfordshire Research Ethics Committee C (07/H0606/148) approved this study. Normal pregnancy was a healthy singleton pregnancy with normal maternal blood pressure. PE was defined as new (after 20 weeks) systolic blood pressure 140 mmHg or diastolic pressure 90 mmHg, proteinuria (protein/creatinine ratio of 30 mg/mmol or more). None of our patients had maternal acute kidney injury, liver dysfunction, neurological features, haemolysis, thrombocytopenia, and/or foetal growth restriction. This study included only early-onset PE patients (diagnosed before 34 weeks gestation). After informed consent, placenta was obtained from women undergoing elective caesarian section without labor were collected.

### Placenta sample preparation and syncytiotrophoblast membrane extracellular vesicles (STB-EVs) enrichment

Placenta biopsies were obtained adjacent to the perfused lobe and immediately frozen at -80°C pending transfer to the Target Discovery Institute (Oxford) for proteomic analysis. We obtained STB-EVs via placental perfusion as previously described(Dragovic et al., 2015b). Our STB-EV enrichment and categorization process has been deposited on EV Track ([http://www.EVTRACK.org], **EV-TRACK ID: EV 220157**) with a score of 78% (the average score on EV track for 2021 is 52 %). Full details can be found in the supplemental data.

### Characterization of syncytiotrophoblast membrane extracellular vesicles (STB-EVs)

Enriched STB-EVs were resuspended in filtered phosphate buffered saline (fPBS) and characterized with bicinchoninic acid (BCA) assay (for protein concentration) and nanoparticle tracking analysis (NTA) (for particle number and size profile). We also phenotyped the STB-EVs with transmission electron microscopy (for morphology), flow cytometry (BD Biosciences, LSRII), and western blot (for immunophenotyping). Flow cytometric analysis was performed using antibodies to placental alkaline phosphatase – PLAP-(to confirm syncytiotrophoblast origin), CD 41 (to identify co-isolated platelet EVs, CD235 a/b (to identify co-isolated red blood cell EVs), HLA class I and II (to identify co-isolated white blood cell EVs). Western blot was probed for placental alkaline phosphatase (PLAP [1.667 mg/ml] at 1:1000 dilution in house antibody), the known EV markers CD 63 ([200ug/ml] at 1:1000 dilution, Sc-59286, Santa Cruz Biotechnology), ALIX ([200ug/ml] at 1:1000 dilution, Sc-53538, Santa Cruz Biotechnology) and the known negative EV marker Cytochrome C ([200ug/ml] at 1:1000 dilution, Sc-13156, Santa Cruz Biotechnology) as recommended by the international society for extracellular vesicles (ISEV) and subsequently incubated with the corresponding secondary antibody anti-mouse, or anti-rabbit polyclonal goat immunoglobulins/HRP (at 1: 2000 dilution, Dako UK Ltd, Cambridgeshire UK). Details of nanoparticle tracking analysis and transmission electron microscopy can be found in the supplemental data.

### Sample preparation for Mass Spectrometric analysis and bioinformatic analysis of proteomic data from placenta tissue, medium/large, and small STB-EVs

STB-EVs and placental tissue samples were processed for liquid chromatography mass spectrometry (LC-MS) (Target Discovery Institute, Oxford). Briefly, STB-EVs and placental tissue samples (10 µg total protein) were reduced with dithiothreitol (DTT) (final concentration 5 mM) for 60 min at room temperature, then alkylated with iodoacetamide (final concentration 20 mM) for 60 min at room temperature. After precipitation with methanol/chloroform, the protein pellet was resuspended in 6 M urea, and then the urea concentration was reduced to < 1 M with milliQ H2O. Trypsin was added to achieve the final trypsin: protein ratio of 1:50, and the samples were digested overnight at 37°C. According to the manufacturer’s instructions, peptides were purified on Waters C18 Sep-Pak cartridges. The purified peptides were dried down in a speed vac, resuspended in 2% acetonitrile/0.1% trifluoroacetic acid, and diluted 1:20 before injection. Peptides were injected into an LC-MS system comprised of a Dionex ultimate 3000 Nano LC (Liquid Chromatography) and a Thermo Q-Exactive mass spectrometer. Peptides were separated on a 50-cm-long EasySpray column (ES803; Thermo Fisher) with a 75 µm inner diameter and a 60-minute gradient of 2% to 35% acetonitrile in 0.1% formic acid and 5% dimethyl sulfoxide (DMSO) at a flow rate of 250 nl/min. The Top 15 most abundant peaks were fragmented after isolation with a mass window of 1.6 and a resolution of 17,500. The normalized collision energy was 28% (higher collisional dissociation). Raw data was imported and analyzed with Progenesis QI (Waters) using standard settings and manually refined retention time alignment. MS/MS data was searched in Mascot (Matrix Science) against a human database (fused Uniprot/Trembl, 03/2018), with oxidation (Met), deamidation (Gln/Asn) and carbamidomethylation (Cys) fixed as variable modifications. Precursor mass tolerance was set to 10 parts-per-million (ppm) and fragment tolerance to 0.04 Da. Peptide identifications were FDR (False Discovery Rate) adjusted at 1%, and identifications with Mascot score < 20 were discarded.

Raw label-free quantitation (LFQ) data was imported and analyzed in Perseus (Max Planck Institute of Biochemistry). Differential expression was performed by conducting a two-sample independent Student t-test. Multiple testing was corrected via permutation-based FDR (False Discovery Rate) with the default software settings. Proteins were considered differentially expressed if their false discovery rate (FDR) was less than 0.05 and their fold change was greater than or equal to 1 or less than or equal to -1. G: Profiler (https://biit.cs.ut.ee/gprofiler/gost) was used for functional enrichment analysis of the differentially expressed proteins (DEPs). The enriched pathways/terms from the KEGG database and Gene ontology database, gene ontology biological process (GO: BP), gene ontology molecular function (GO: MF), and gene ontology clinical component (GO: CC) were ascertained for each set of DEPs by applying hypergeometric testing. Multiple testing was corrected by Benjamin Hochberg correction, and the significance level was set to < 0.05.The mass spectrometry proteomics data have been deposited to the ProteomeXchange Consortium via the PRIDE partner repository with the dataset identifier PXD031953.132

### Bioinformatic analysis of proteomic data from placenta tissue, medium/large and small STB-EVs

Persus (Max Planck Institute of Biochemistry) was used for the analysis, along with the accompanying documentation and tutorials. To remove invalid data, we pre-processed the raw data by log transforming and filtering. Missing values were imputed at random from a normal distribution using the following parameters: width = 0.3 and downshift = 1.8. To ensure conformity to the normal distribution, the underlying distribution was visually inspected with a histogram before and after missing data imputation. The data was then transformed further by deducting each (transformed) value from the highest occurring protein expression value.

Principal component analysis (PCA), heatmaps, and Pearson correlation matrices were used to further investigate the data. The data was analyzed using the correlation index and hierarchical clustering. A two-sample independent student t-test was used to assess differential expression. Multiple testing was corrected using permutation-based false discovery rate (FDR) with the following parameters, with significance set at less than 0.05.

### Western Blotting

To further characterize and immune-phenotype, we performed western blots on placental homogenates and STB-EV pellets. The relevant primary antibodies were used to probe all STB-EVs (as discussed in the main article). An equal amount of protein (20 micrograms) was mixed with 4 X Laemmli buffer ((180 mM Tris-Cl (pH 6.8), 6% SDS, 30% glycerol, 0.3% 2-mercaptoethanol, and 0.0015% bromophenol blue, BioRad)). After heating the sample mix for ten minutes at 70°C, an equal volume of the sample mix was loaded. Electrophoresis was performed at 150 V for 1.5 hours in a Novex minicell tank (Invitrogen, UK) filled with NuPAGE^TM^ MOPS SDS running buffer under reducing (for PLAP, Cytochrome C, ALIX, SR-BI, Filamin B, PAPP-A2, Collagen 17 A1) and non-reducing (for CD 63) conditions on NuPAGE TM 4-12% Bis-Tris Gel 1.0 mm x 10 well gels (Invitrogen by Thermo Fisher Scientific) (Novex by Life Technologies). As a protein size marker, Precision plus protein^TM^ dual color standards (Bio-Rad Laboratories Ltd, Hertfordshire, UK) were used. Following protein separation, the proteins were transferred to a polyvinylidene difluoride (PVDF) membrane (Bio-Rad).

In a Novex semi-dry transfer apparatus, the PVDF membrane and gel were sandwiched between four filter paper sheets pre-soaked in anode one buffer solution (300 mM Tris, 10% methanol pH 10.4) and two filter paper sheets pre-saturated with anode two buffer solution (25 mM Tris, 20% methanol pH 10.4) at the bottom and three filter paper sheets pre-soaked in cathode buffer solution (25 mM Tris, 40 mM (Life Technologies, UK). For 45 minutes, the transfer was run at 25 V. The membranes were blocked for one hour with 5% Blotto (2BScientific) in 0.1% TBST (Tris-buffered saline (20 mM Tris, 137 mM NaCl, pH 7.6) and 1% Tween-20, Sigma), then incubated overnight with the appropriate antibody.

Antibodies used were SR-BI (ab52629 [0.35 µg/µl] 1:1000 dilution, monoclonal Rabbit, Abcam), Filamin B (GTX387 [1.64 µg/µl] 1: 1000 dilution, monoclonal mouse, Insight Biotechnology), PAPP-A2 (ab59100 [1 µg/µl] 1: 2000 dilution polyclonal Rabbit, Abcam), Collagen 17 A1(ab28440 [1 µg/µl] 1: 2000 dilution polyclonal Rabbit, Abcam), and for characterization, placental alkaline phosphatase (PLAP [1.667 µg/µl] at 1:1000 dilution in house antibody), CD 63 ([200 µg/µl]] at 1:1000 dilution, Sc-59286, Santa Cruz Biotechnology), Alix ([200µg/µl]] at 1:1000 dilution, Sc-53538, Santa Cruz Biotechnology), and Cytochrome C ([200 µg/µl]] at 1:1000 dilution, Sc-13156, Santa Cruz Biotechnology) as recommended by the international society for extracellular Vesicles (ISEV).

After an overnight incubation and three five-minute TBST washes, the membranes were incubated for an hour at room temperature with the corresponding secondary antibody, anti-mouse (P044701-2) or anti-rabbit (P044801-2) polyclonal goat immunoglobulins/ horseradish peroxidase (HRP) (1: 2000 dilution, Dako UK Ltd, Cambridgeshire UK) and rinsed three times in TBST. The membranes were developed using a gel documentation system (G-Box, Syngene, Cambridge UK) running GeneSys (version 1.5.0.0, Syngene) and a chemiluminescence film (Amersham HyperfilmTM ECL (enhanced chemiluminescence; GE Healthcare Limited, Buckinghamshire UK) to obtain the band intensity for each lane. The fold change (FC) between normal and preeclampsia samples was calculated by normalizing the comparative expression analysis to the total protein loaded (using Amido Black stain) 17,18. The normalized densitometric values were statistically tested using a one-tailed Student t-test, with significance set at less than 0.05. Tables 1 and 2 show the reagents and antibodies used in western blot analysis, respectively. Details of western blot for STB-EV characterization and proteomic validation experiments can be seen in the supplemental material.

**Table 1.** General descriptive statistics of sample population.

| Characteristics | Sub-Classification | Normal Pregnancy | Preeclampsia | P Value |
| --- | --- | --- | --- | --- |
| Sample size |  | 6 | 6 |  |
| Maternal age years (mean (SD)) |  | 34.5 ± 5.39 | 36.33 ± 4.13 | 0.524 |
| Systolic blood pressure mmHg (mean (SD)) |  | 129.5 ± 4.93 | 178.83 ± 12.56 | <0.001 |
| Diastolic blood pressure mmHg (mean (SD)) |  | 67 ± 6.2 | 109.17 ± 9.85 | <0.001 |
| Body mass index kg/m-2 (mean (SD)) |  | 29.92 ± 9.1 | 31.25 ± 11.12 | 0.825 |
| Proteinuria plus(es) (mean (SD)) |  | 0 | 2.58 ± 1.2 | <0.001 |
| Gestational age at diagnosis in weeks (mean (SD)) |  | NA | 30.5 ± 3.50 | 0.001 |
| Gestational age at delivery in weeks (mean (SD)) |  | 39.17 ± 0.98 | 32 ± 3.52 | 0.001 |
| Birth weight grams (range) |  | 3912.5 ± 730.4 | 1515.83 ± 600.57 | <0.001 |
| Intrauterine growth restriction (IUGR) = Yes (%) |  | 0(0) | 6(100) | 0.004 |
| Male new-born sex (%) |  | 2(33.3) | 2(33.3) | 1.000 |

This study utilized 12 samples for initial discovery and 12 for targeted western blot validation

## Results

### Patient demographics and clinical characteristics

PE mothers (Table 1), as expected, had a significantly higher average systolic (178.83 mmHg, P < 0.001) and diastolic (109.17 mmHg, P < 0.001) blood pressure compared to normal pregnant mothers average systolic (129.50 mmHg) and diastolic (67.00 mmHg). PE women are also significantly more likely to deliver prematurely (PE = 32.00 weeks gestation NP = 39.17 weeks gestation, P < 0.001) and have proteinuria (PE = 2.58 pluses on urine dipstick NP = 0 pluses on urine dipstick, P < 0.001) compared to normal pregnancy Finally, PE babies significantly weighed less than normal babies (PE = 1515.83g NP = 3912.50 g, P < 0.001). Surprisingly, we found no significant difference in body mass index, the gender of the child, and maternal age.

### Characterization of syncytiotrophoblast membrane extracellular vesicles (STB-EVs)

Flow cytometry (Figure 1) was showed that many detected events (83-85 ± 8.0-8.3%) were negative for CD 235a (Red blood cells), CD41 (platelets) and HLA-I and II (white blood cells) (Figure 1B) while 92 ± 0.9% (Figure 1D and 1G) of detected events were PLAP^+^ extracellular vesicles (BODIPY FL N-(2-aminoethyl)-maleimide (bioM) and placental alkaline phosphatase (PLAP) double-positive). Detergent treatment, which could break down EVs, with NP-40 confirmed that majority (99%) of our samples were largely vesicular since only 0.1 ± 0.12% of BODIPY FL N-(2-aminoethyl)-maleimide and PLAP double-positive events were detected (a reduction of 99%) (Figure 1E and 1H).

**Figure 1.**
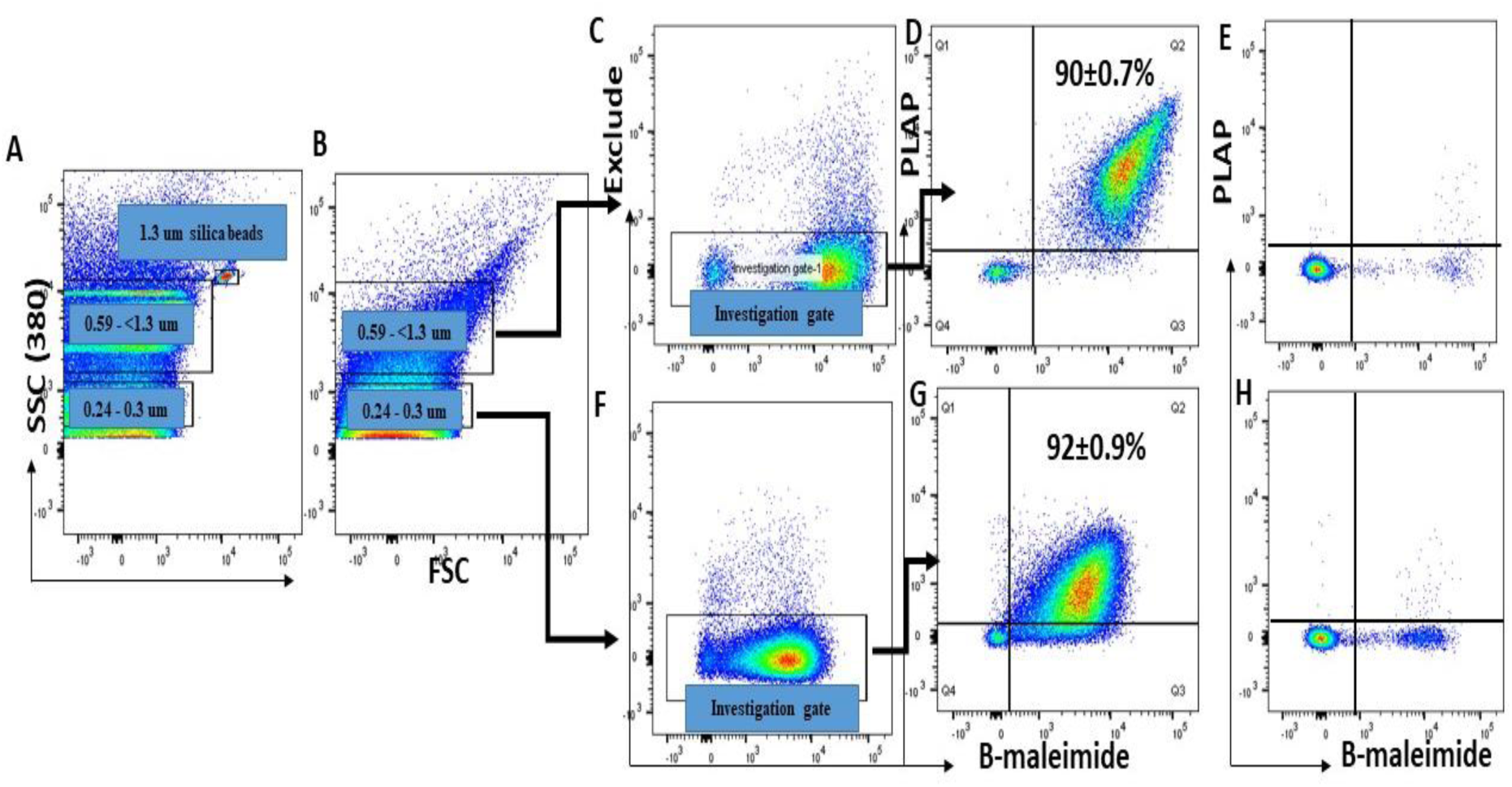
Flow analysis of medium/large STB-EVs in the 10K STB-EV pellet. Apogee beads mix were used to set the flow machine’s light scatter resolution to 0.59-1.3 µm and 0.24-1.3 µm silica beads(A). Figure B shows the application of SSC and FSC PMTVs as determined by apogee beads mix for the analysis of m/lSTB-EVs in the 10K pellet. An investigation gate was created to include only medium/large EVs negative for non-placental markers (C & F). Extracellular vesicles from the investigation gate were further analyzed for staining by bioM and expression of PLAP (D & G). Figure E and H shows that the bioM^+^ PLAP^+^ EVs were sensitive to detergent treatment. The percent of B-Maleimide^+^ PLAP^+^ from the 0.59-1.3 µm gate is consistent under both SSC conditions.

Transmission electron microscopy on 10K STB-EV pellets (Figure 2A and 2B) and 150K STB-EVs (Figure 2C and 2D) in our sample preparation showed the typical cup-shaped morphology of extracellular vesicles on transmission electron microscopy (TEM) within the appropriate size range. Western blot confirmed they express the classic syncytiotrophoblast membrane marker, placenta alkaline phosphatase (PLAP), the extracellular vesicle markers ALIX and CD 63. In addition, they lacked the negative EV marker cytochrome C (Figure 3A). Nanoparticle tracking analysis (NTA) confirmed the homogeneity of the 150K STB-EV pellets (small STB-EVs) (Figure 3B) with a modal size of (205.8 ± 67.7) nm and the heterogeneity of the 10K STB-EV pellets (medium/large STB-EVs) (Figure 3C) with a size range of (479.4 ± 145.6) nm.

**Figure 2.**
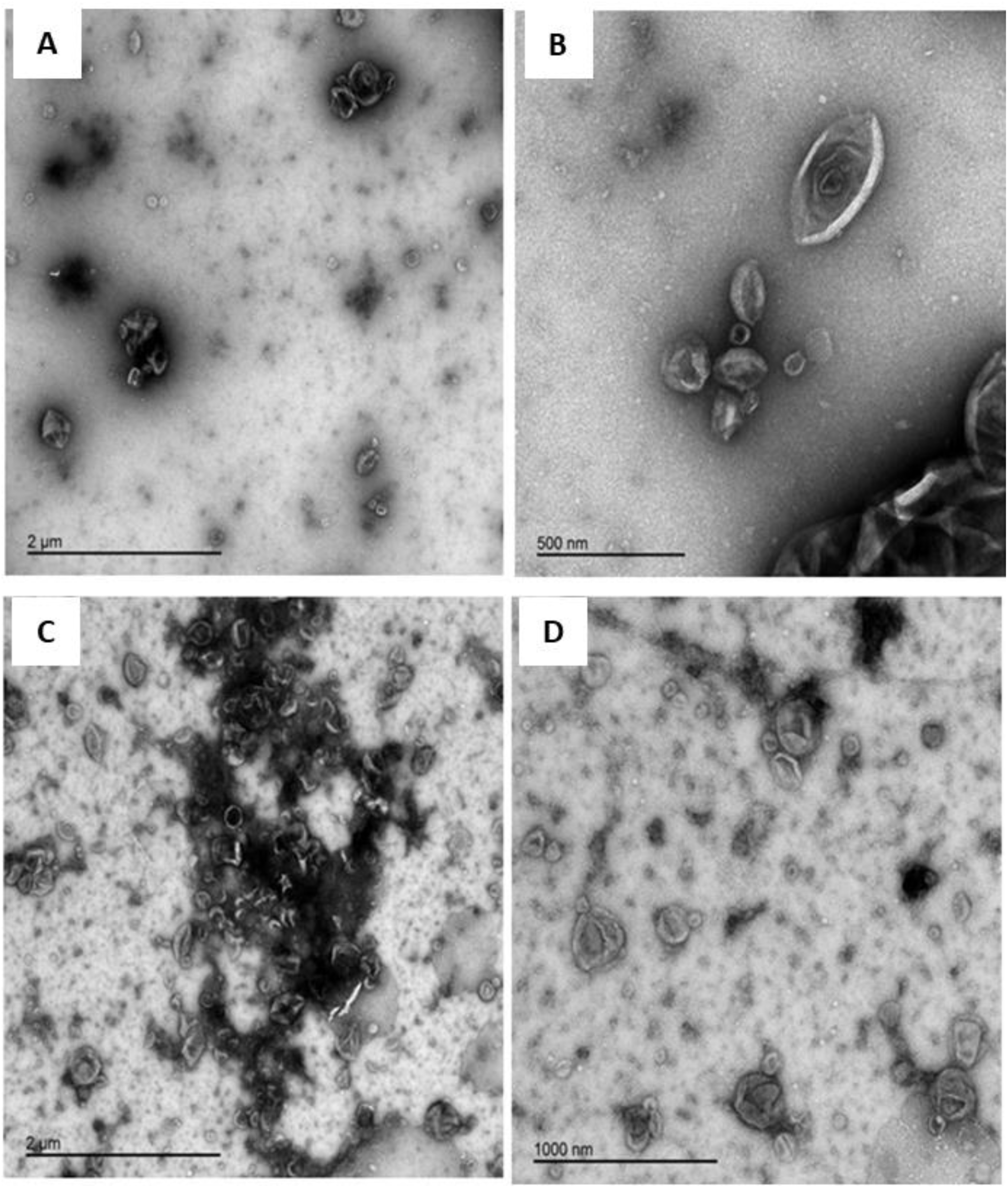
Results of STB-EV characterization. Figure A,B,C and D displays representative transmission electron microscopy (TEM) images with wide view (A and C), medium/large STB-EVs (B), and Small STB-EVs (D).

**Figure 3.**
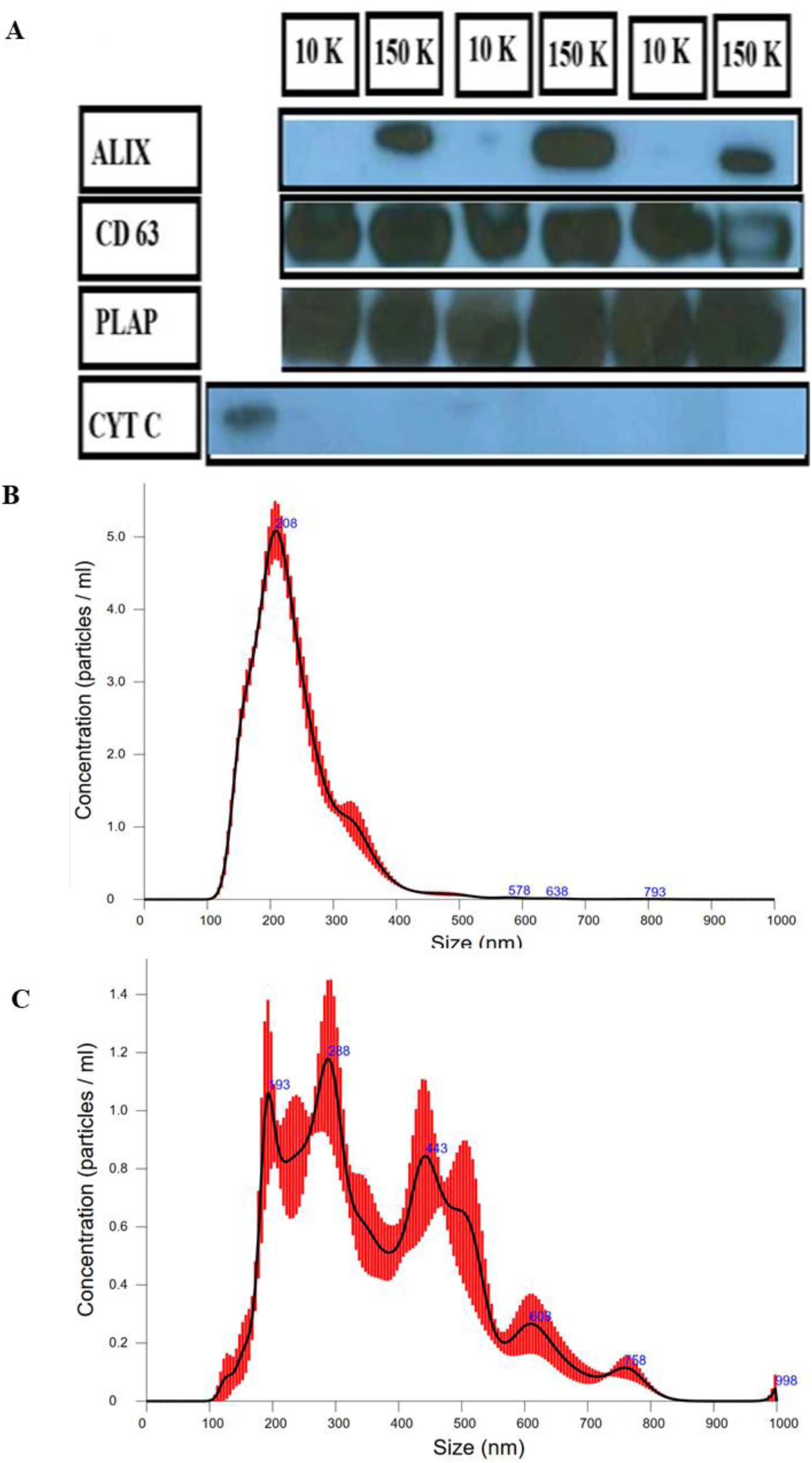
Results of STB-EV characterization. The western blot characterization of S STB-EVs and mlSTB-EVs (A). 10K refers to mlSTB-EVs and 150K refers to s STB-EVs. CYT C refers to cytochrome c. Figure 2A and 2B show the NTA results of m/lSTB-EVs (B) and sSTB-EVs (C).

### Differentially carried proteins (DCPs) in Placenta homogenate, Medium/Large STB-EVs and Small STB-EVs in Preeclampsia versus normal pregnancies

In total, using mass spectrometry, there were fifteen (15) proteins in the placenta, three hundred and four (304) in m/lSTB-EVs, and seventy-three (73) in sSTB-EVs were differentially expressed between preeclampsia (PE) and normal pregnancy (NP).

In the placenta (Table 2)*, isoform HMG-R of High mobility group protein (HMGA1), Fibrinogen-like protein 1(FGL1), isoform 1 of Kinesin-like protein (KIF2A), Ig kappa chain C region (IGKC)* were the most abundant proteins based on fold change. Concomitantly, *serum paraoxonase/arylesterase 1 (PON1) and alpha-1B-glycoprotein (AIBG)* were the least abundant proteins.

**Table 2:** Top ten differentially expressed Proteins between normal and preeclampsia placentas (bold font), medium/large STB-EVs (normal font), small STB-EVs(italics)

| Protein | Symbol | LFC | Adjusted P. Value |
| --- | --- | --- | --- |
| Placenta |  |  |  |
| **Isoform HMG-R of High mobility group protein HMG-I/HMG-Y** | **HMGA1** | **3.14** | **0.03** |
| **Fibrinogen-like protein 1** | **FGL1** | **2.49** | **0.04** |
| **Isoform 1 of Kinesin-like protein KIF2A** | **KIF2A** | **1.47** | **0.02** |
| **Chloride intracellular channel protein 3** | **CLIC3** | **1.09** | **0.05** |
| **Mesencephalic astrocyte-derived neurotrophic factor** | **MANF** | **1.02** | **0.03** |
| **Ig kappa chain V-I region AG** | **KV101** | **-1.02** | **0.05** |
| **Haloacid dehalogenase-like hydrolase domain-containing protein 2 (Fragment)** | **K7EJQ8** | **-1.23** | **0.05** |
| **Ig kappa chain C region** | **IGKC** | **-1.27** | **0.05** |
| **Alpha-1B-glycoprotein** | **A1BG** | **-1.34** | **0.05** |
| **Serum paraoxonase/arylesterase 1** | **PON1** | **-2.18** | **<0.01** |
| Medium/Large STB-EVs |  |  |  |
| Collagen alpha-1(XVII) chain | COL17A1 | 4.79 | <0.01 |
| Isoform 2 of Filamin B | FLNB | 3.09 | <0.01 |
| Tumor necrosis factor alpha-inducible protein | TNFAIP2 | 3.03 | <0.01 |
| Minor histocompatibility antigen | HMHA1 | 1.59 | 0.03 |
| Bridging integrator 2 | BIN2 | 2.33 | 0.02 |
| Sodium/potassium-transporting ATPase subunit beta-1 | ATP1B | 1.99 | 0.02 |
| ERO-1 like protein alpha | ERO1L | 1.97 | <0.01 |
| Endoglin | ENG | 1.82 | <0.01 |
| Methylthioribose-1-phosphate isomerase (Fragment) | MRI1 | -1.96 | 0.04 |
| Sodium-dependent phosphate transporter 1 | S20A1/SLC17A1 | -5.24 | 0.04 |
| Small STB-EVs |  |  |  |
| *Solute carrier family 2, facilitated glucose transporter member 11* | *SLC2A11* | *8.91* | *0.01* |
| *V-type proton ATPase 16 kDa proteolipid subunit* | *VATL* | *7.25* | *0.02* |
| *Pappalysin-2* | *PAPP2* | *3.76* | *0.02* |
| *ERO1-like protein alpha* | *ERO1A* | *3.29* | *0.02* |
| *Isoform 2 of Creatine kinase U-type, mitochondrial* | *KCRU* | *3.20* | *0.02* |
| *SCARB1 protein* | *B7ZKQ9* | *2.95* | *0.05* |
| *Isoform 2 of Filamin-B* | *FLNB* | *2.92* | *0.02* |
| *Solute carrier organic anion transporter family member 2A1* | *SO2A1* | *2.86* | *0.02* |
| *Isoform 2 of Oligaccharyltransferase complex subunit TC* | *OSTC* | *2.82* | *0.02* |
| *SCY1-like protein 2* | *SCYL2* | *2.71* | *0.02* |
| *Sodium-dependent phosphate transporter 1* | *S20A1* | *-2.93* | *0.01* |

For m/lSTB-EVs (Table 2), the most differentially abundant proteins were the *collagen alpha-1(XVII) chain (COL17A1), isoform 2 of Filamin-B (FLNB), tumor necrosis factor-alpha-induced protein 2 (Fragment) (TNFAIP2)* based on fold change. In contrast, the least differentially abundant proteins were *sodium-dependent phosphate transporter 1 (SLC20A1), methylthioribose-1-phosphate isomerase (Fragment) (MRI1), prostatic acid phosphatase (Fragment) (ACPP*) based on fold change.

Finally, the sSTB-EVs (Table 2) had *solute carrier family 2, facilitated glucose transporter member (SLC2A11), v-type proton ATPase 16 KDa proteolipid subunit (ATP6V0C), pappalysin 2 (PAPP-A2)* as the most differentially abundant by fold change while *sodium-dependent phosphate transporter 1 (SLC20A1)* and *isoform 2 of ADP-ribosylation factor GTPase-activating protein (ARFGAP3)* were the least differentially abundant. There were 25 differentially carried proteins (DCPs) including *filamin B (FLNB), ERO1-like protein alpha (ERO1A), endoglin (EGLN), pappalysin-2 (PAPP-A2), siglec6 (SIGL6)* shared between m/lSTB-EVS and sSTB-EVs and only one protein, *isoform 1 of kinesin-like protein (KIF2A)* shared between the placenta and the m/l STB-EVs. Only one protein *chloride intracellular channel protein 3 (CLIC3),* was found in all three sample sub-types (Table 3)

**Table 3.** List of overlapping differentially carried proteins in the placenta, medium/large STB-EVs and small STB-EVs.

| Sample types | Differentially expressed proteins (DEPs) |
| --- | --- |
| m/ISTB-EVs and Placenta | Kinesin-like protein |
| m/ISTB-EVs, sSTB-EVs and Placenta | Chloride intracellular channel protein 3 |
| m/ISTB-EVs and sSTB-EVs | Filamin B |
|  | Endoplasmic Reticulum Oxidoreductase Alpha |
|  | X-linked retinitis pigmentosa GTPase regulator interacting protein 1 |
|  | Protein NDRG1 |
|  | Collagen alpha 1(XVII) chain |
|  | Monocyte differentiation antigen CD14 |
|  | Trophoblast glycoprotein |
|  | Heat shock-related 70 kDa protein 2 |
|  | Endoglin |
|  | Hippocalcin-like protein 1 |
|  | SCARB1 protein |
|  | Importin subunit alpha 7 |
|  | Protein BCAP |
|  | Endothelial protein C receptor |
|  | Caveolin |
|  | Pappalysin-2 |
|  | Phosphatidylinositol 3-kinase regulatory subunit alpha |
|  | Keratin, type I cytoskeletal 19 |
|  | Siglec6 |
|  | Protein disulfide-isomerase |
|  | Amine oxidase [flavin-containing] A |
|  | Copine-3 |
|  | Dolichyl-diphosphooligosaccharide |
|  | Sodium-dependent phosphate transporter 1 |
|  | Annexin A4 |

### Validation of select proteins in the placenta homogenate, medium/Large STB-EVs, and small STB-EVs

We combined all the DCPs identified from the placenta, m/lSTB-EVs, and sSTB-EVs and selected proteins to validate based on fold change and our placenta specificity or enrichment as previously described above in the methods section. These were pappalysin A2 (*PAPP-A2), collagen 17A1 (COL17A1), filamin B,* and *scavenger receptor class B type I (SR-BI/ SCARB1)*. Validation was performed by western blot on all sample sub-types.

In the placenta (Figure 4A and 4D), *PAPP-A2* (FC =6.39, P Value = 0.0001) and *SR-BI* (FC = 1.72, P Value = 0.04) were all differentially abundant in PE. *Filamin B* (FC = 2.59, P Value = 0.12) was differentially abundant in PE but not significant while *COL17A1* was undetectable. In medium/large STB-EVs (Figure 4B and 4E), pappalysin A2 (FC =5.09, P Value = 0.004), *collagen 17A1* (FC =71.21, P Value = 0.002), *filamin B* (FC =7.60, P Value = 0.014) and *scavenger receptor class B type I* (FC =2.28, P Value = 0.018) were all significantly differentially abundant in PE compared to normal. In sSTB-EVs (Figure 4C and 4F), only *filamin B* (FC =7.73, P Value = 0.003) and *SR-BI/SCARB1* (FC =1.60, P Value = 0.002) were significantly differentially abundant while *COL17A1*(FC =12.79, P Value = 0.07) and *PAPP-A2* (FC =1.48, P Value = 0.11) were differentially abundant but not significantly so.

**Figure 4.**
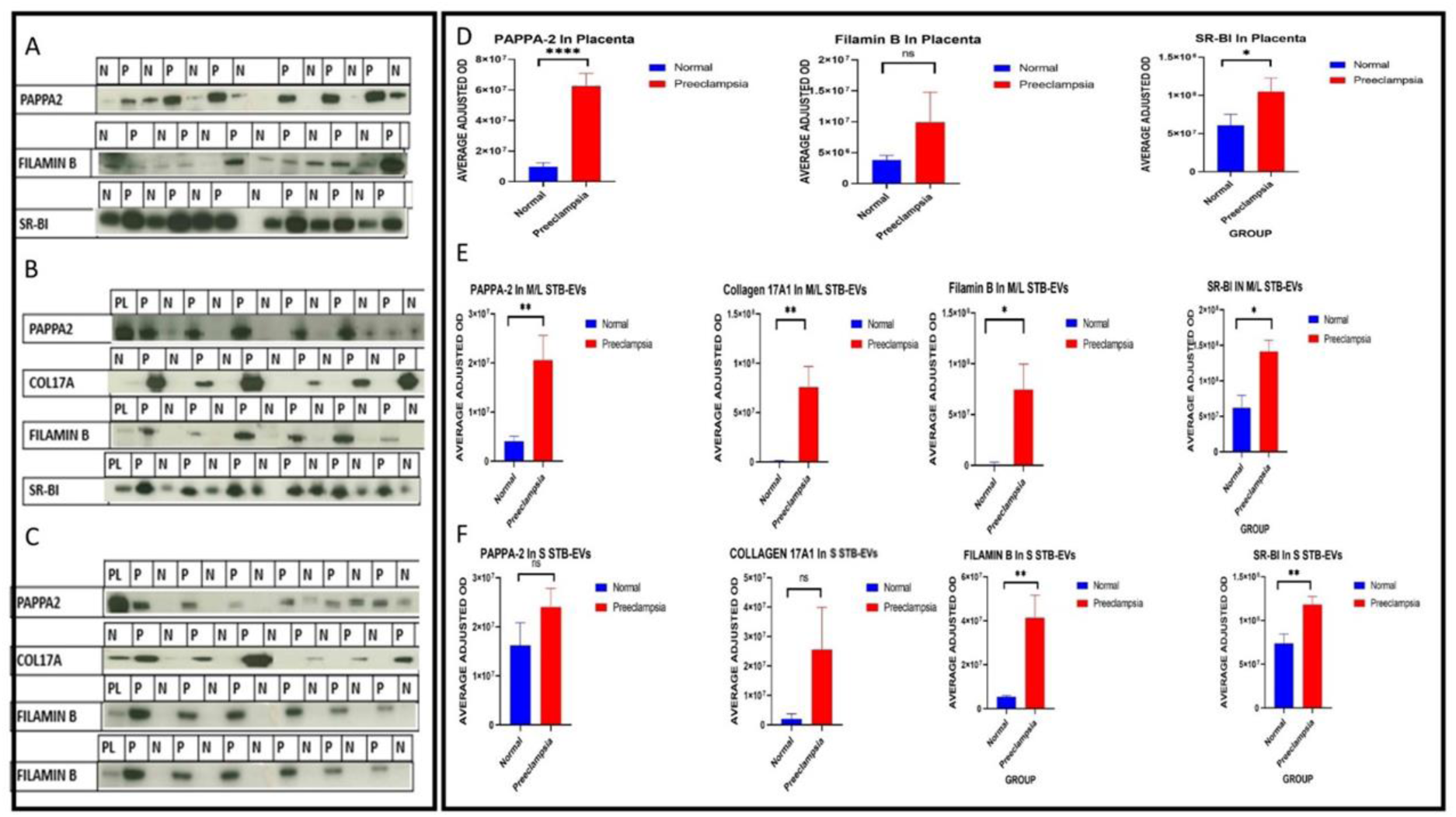
Figure showing the results of western blot and densitometric verification of PAPP-A2, COL17A1, FLNB and SR-BI in the placenta (A and D) m/lSTB-EVs (B and E) and sSTB-EVs (C and F). Collagen 17A1 did not show up in the placenta homogenate. * means < 0.05, ** means < 0.01, *** means < 0.001, **** means less than 0.001 and ns means non-significant.

### Functional enrichment of differentially carried proteins (DCPs) in preeclampsia (PE)

We performed a functional enrichment analysis on the list of differentially carried proteins in placenta tissue, medium/large STB-EVs, and small STB-EVs to help better understand their role in preeclampsia (PE). The three sample sub-types did not have overlapping gene ontology terms or KEGG pathways. The top biological processes overrepresented in the placenta (Table 3A) were processes that involve *neurotransmitter secretion and transport*. In comparison, the enriched biological processes in the m/lSTB-EVs (supplemental table 3) *involved responses to hypoxia*. Finally, *posttranslational protein modification processes* were enriched among small STB-EVs (supplemental table 3). *Neurotrophin signaling pathway, spinocerebellar ataxia, and protein processing pathway* were among the over enriched KEGG pathways in the placenta, m/lSTB-EVs and sSTB-EVs respectively (Supplemental table 4).

## Discussion

### Principal findings

In our analysis, PE and NP have different placenta and STB-EV proteomes. Four STB-EV biomarkers—filamin B, collagen 17A1, pappalysin-A2, and scavenger Receptor Class B Type 1—were verified for differential abundance. In silico investigation revealed molecular pathways such abnormal protein metabolism that may contribute to PE’s clinical and pathological symptoms and inform future research.

### Results in the context of what is known

Our analysis found chloride intracellular channel 3 (CLIC3) to be the only protein differentially abundant among all sample types. CLIC3 is expressed in the placenta throughout pregnancy(Money et al., 2007) and is abundant in PE placentas compared to NP, a finding corroborated by our results. However, it has not previously been described in terms of STB-EVs. CLIC3 also recycles activated integrins back to the plasma membrane and facilitates cell migration and invasion(Dozynkiewicz et al., 2012) a process that has been identified as abnormal in PE(KHONG et al., 1986).

We found *COL17A1* to be more abundant in PE m/lSTB-EVs but not detectable in the placenta in both normal and PE. Although collagen 17 has not been described in preeclampsia, a protein of the same family, collagen 1 is deposited in higher amounts in the PE placenta and can induce preeclampsia-like symptoms by suppressing the proliferation and invasion of trophoblasts. This suppression was reversible by treating with ERK and B-catenin agonists(Feng et al., 2021). Likewise, *PAPP-A2* was significantly more abundant in the PE placenta and m/lSTB-EVs. *PAPP-A2* cleaves insulin-like growth factor binding protein (IGFBP-5 and, to a lesser extent, IGFBP-3). PAPP-A2’s mRNA and protein are differentially expressed in the placenta and maternal serum in PE patients(Whitehead et al., 2013).

Filamin B was significantly more abundant in PE m/lSTB-EVs and sSTB-EVs compared to NP. Filamin B participates in cellular structural mechanics and signal transduction by interacting with ion channels, signaling molecules, transmembrane proteins, and transcription factors(Zhou et al., 2010). It also suppresses tumor growth and metastasis(Iguchi et al., 2015). Interestingly, in contrast to our study wei et al described Filamin B as being up regulated in the placenta (Wei et al., 2019) . Wei et al used glyceraldehyde 3-phosphate dehydrogenase (GAPDH) as an internal reference while we used total protein normalization to Amido black to quantify and compare protein expression. GAPDH is an unreliable internal reference protein, particularly in preeclampsia(Lanoix et al., 2012) and this may explain these discordant findings. In addition, in our proteomics data, we found GAPDH to be among the most differentially expressed (upregulated) proteins.

Likewise, we identified scavenger receptor class B, type 1 (SCARB1/SR-BI) to be significantly increased in PE placenta, m/l and sSTB-EVs. SCARB1/SR-BI, is most abundant in the adrenal glands, placenta, liver, and brain(Ganesan et al., 2016; Shen et al., 2016). SR-BI facilitates the uptake of cholesteryl esters from high-density lipoproteins and lipid-soluble vitamin and transthyretin-bound thyroid hormone by placental trophoblast cells(Landers et al., 2018).

In terms of potential mechanisms of preeclampsia, we found no overlap in the biological processes and KEGG pathways among the three sample sub-type. In the placenta, gene ontology biological processes (GO: BP) involved in *neurotransmitter secretion and transport* were overrepresented while *platelet activation*, *MAPK* and *Rap 1 signaling* pathways were among the detected KEGG pathways. In m/lSTB-EVs, *protein modification process,* and *decreased oxygen level responses* were among the perturbed GO: BP terms. *Alzheimer’s* and *prion disease* were among the identified KEGG pathways. In sSTB-EVs, *post-translational modification processes* and *endoplasmic reticulum protein processing* were the principal GO: BP terms and KEGG pathways.

Recent research has shown that ischemic hypoxia and the release of proinflammatory cytokines in PE can cause protein misfolding and initiate endoplasmic reticulum (ER) stress due to hypoxia-reoxygenation damage to the endoplasmic reticulum(Gathiram & Moodley, 2016). PE can also cause posttranslational modifications to proteins, such as changing the isoelectric point, which results in different S-nitrosylation outcomes in placental proteins(Zhang et al., 2011). It is thought that the accumulation of these aggregates of unfolded protein response (UPR) or misfolded proteins contributes to the pathophysiology of PE(Gathiram & Moodley, 2016). Other processes and KEGG pathways have been previously described in preeclampsia, while others found in our study are new and may warrant further research(Lee et al., 2020; Wan Shumei, Peng Ping, Qiao Lin, 2019).

### Ideas and Speculation

STB-EVs are liquid biomarkers with real-time information from the damaged placenta in PE due to their placenta-specificity. It would be interesting to test maternal plasma or serum samples for the four STB-EV indicators reported in our study.

It is unclear which comes first: misfolded proteins depositing in trophoblasts and preventing normal invasion, causing ischemia and endoplasmic reticulum stress, which leads to defects in trophoblast invasion, oxidative stress, and endothelial cell dysfunction, or the trophoblast invasion defects, oxidative stress, and endothelial cell dysfunction, or the misfolded proteins because of faulty invasion and oxidative stress. Further studies exploring these PE pathogenic processes would be intriguing.

### Strengths and limitations

Our study explored the difference in the proteome between PE and NP by analyzing the placenta and its extracellular vesicles. This study is one of the few to do so using extracellular vesicles obtained by a physiologic technique, the *ex-vivo* dual lobe placenta perfusion. However, the control population used for this study was not gestationally age-matched because it is impossible to obtain the ideal control for early onset PE patients. Also, our sample size was small (n of 12).

## Conclusion

Our study may have found novel STB-EV-bound protein indicators that are significantly more abundant in PE than normal. Since STB-EVs are present in the circulation from early pregnancy to term and are released more in PE, these STB-EV carried proteins may help with earlier diagnosis and mechanistic insights.

## Data Availability

All data produced are available online at https://data.mendeley.com/datasets/j2x4h9ddcj/draft?a=8ccd383c-c6e5-495d-b66b-29c691608995

## Acknowledgments

We acknowledge the support of the National Institute of Health Research Clinical Research Network for assistance in patient recruitment and Fenella Roseman and Lotoyah Carty, research midwives who kindly recruited the patients for this study. We acknowledge the patients who kindly consented to this research study.

## Supplemental Material

Supplemental Methods

Supplemental table S1-4

Supplemental figure S1

List of differentially abundant proteins and functional enrichment analysis: https://data.mendeley.com/datasets/j2x4h9ddcj/draft?a=8ccd383c-c6e5-495d-b66b-29c691608995

## Supplemental data

### Supplemental methods

**Supplemental figure 1.**
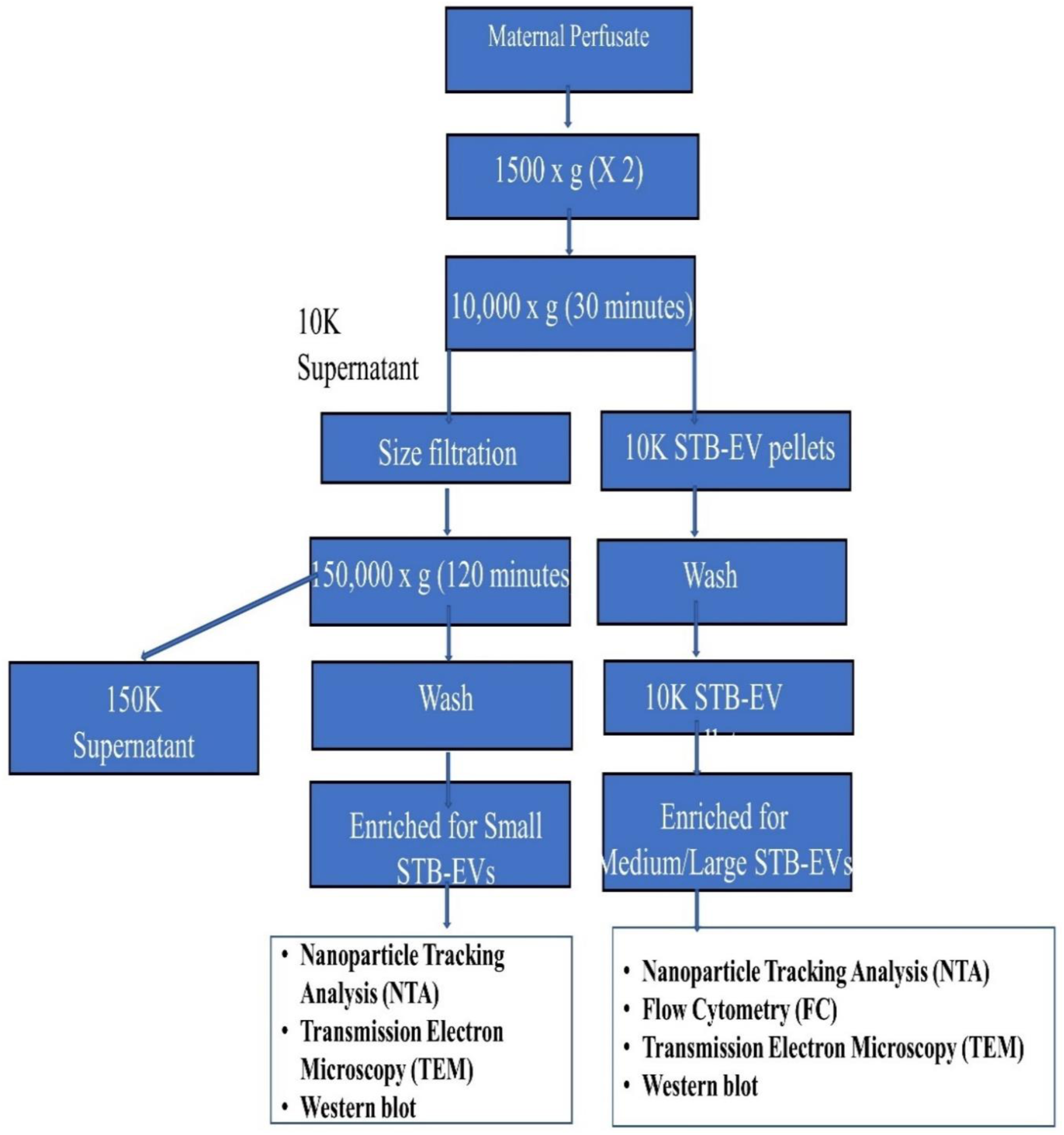
Flow chart illustrating the steps involved in characterizing m/lSTB-EVs and sSTB-EVs obtained via differential ultracentrifugation (10,000 and 150,000 g) of maternal perfusate. Nanoparticle trafficking analysis, transmission electron microscopy, flow cytometry, and western blot were used to characterize the pellets from both spins.

### Transmission electron microscopy

The Sir William Dunn School of Pathology was contracted to perform transmission electron microscopy. STB-EV pellets were diluted with fPBS to produce STB-EV solutions with concentrations ranging from 0.1 to 0.3 g/l. For 2 minutes, ten microliter of the STB-EV pellet solution was applied to freshly glowing discharged carbon formvar 300 mesh copper grids, blotted with filter paper, stained with 2% uranyl acetate for 10 seconds, blotted, and air-dried. The grid’s STB-EV pellets are negatively stained to increase the contrast between the STB-EV pellets and the background. The grids were imaged with a Gatan OneView CMOS camera on an FEI Tecnai 12 TEM at 120 kV.

### Nanoparticle tracking analysis

The Nanosight NS500 (instrument equipped with a 405 nm laser [Malvern UK]), sCMOS camera, and nanoparticle tracking analysis (NTA) software version 2.3, Build 0033 (Malvern UK)) system wased used for the analysis. Instrument performance was tested with silica 100 nm microspheres prior to sample analysis (Polysciences, Inc.). The samples were diluted in fPBS to a concentration to 1/100,000 based on the starting concentration. The samples were automatically injected into the sample chamber with a 1 ml syringe using the EV measurement script: prime, delay of 5, capture of 60, and repeat of 4. Camera images of the analyzed samples were captured at a level of 12. (Camera shutter speed; 15 ms and Camera gain; 350). NTA post-acquisition settings were optimized and maintained constant across samples. Each video recording was analyzed to determine the size and concentration profile of STB-EV.

### Western blotting

**Supplemental table 1.** Reagents used for western blot.

| Buffers | Power of hydrogen (pH) | Constituents |
| --- | --- | --- |
| 4X Laemmli reducing buffer | 6.8 | 9 parts 4 X Laemmli buffer |
|  |  | 1 part 2-beta-mercaptoethanol |
| 4X Laemmli non-reducing buffer |  | 4 X Laemmli buffer |
| Anode buffer 1 | 10.4 | 300 mM Tris base (36.34 g/L) |
|  |  | 100 % methanol (20 mL/L) |
|  |  | 980 ml of ddH<sub>2</sub>O |
| Anode buffer 2 | 10.4 | 25 mM Tris base (3.0 g/L) |
|  |  | 100 % methanol (20 mL/L) |
|  |  | 980 ml of ddH<sub>2</sub>O |
| Cathode buffer | 9.4 | 25 mM Tris base (3.0 g/L) |
|  |  | 40 nM 6-aminocaproic acid (5.2 g/L) |
|  |  | 100 % methanol (20 mL/L) |
|  |  | 980 mL of dH<sub>2</sub>O |
| 10 X TBS | 8 | NaCl (87.6g) |
|  |  | Tris base (12.1g) |
|  |  | 700 ml of ddH<sub>2</sub>O |
| TBST | 8 | 10 X TBS (100 mL) |
|  |  | 0.1 % Tween-20 (1 mL) |
|  |  | 900 mL of ddH<sub>2</sub>O |
| 5% Milk TBST |  | 5g of Blotto in 100 mL of TBST |

**Supplemental table 2.** Antibodies used for western blot.

| Antibodies | Concentration | Dilution | Antigen | Specificity | Manufacturer |
| --- | --- | --- | --- | --- | --- |
| Anti-PLAP (NDOG 2) | 1.6 µg/µl | 1/1000 | PLAP | STB-EV | In house antibody |
| Anti-CD63 | 200 µg/µl | 1/1000 | CD63 | STB-EV | Santa Cruz Biotechnology |
| Anti-ALIX | 200 µg/µl | 1/1000 | ALIX | S STB-EV | Cell Signalling |
| Anti-Cytochrome C | 200 µg/µl | 1/500 | Cytochrome C | Placenta homogenate | Santa Cruz Biotechnology |
| Anti-SR-BI | 0.35 µg/µl | 1/1000 | SR-BI | N/A | Abcam |
| Anti-Filamin B | 1.64 µg/µl | 1/1000 | Filamin B | N/A | Insight Biotechnology |
| Anti-PAPP-A2 | 1 µg/µl | 1/2000 | PAPP-A2 | N/A | Abcam |
| Anti-Collagen 17 A1 | 1 µg/µl | 1/1000 | Collagen 17 A1 | N/A | Abcam |
| Polyclonal goat-anti-mouse/rabbit immunoglobulin HRP |  | 1/2000 | Mouse and Rabbit Immunoglobulins | N/A | Dako UK Ltd |

### Bioinformatic analysis of proteomic data from placenta tissue, medium/large and small STB-EVs

#### Functional enrichment of differentially expressed proteins (DEPs) in preeclampsia (PE)

**Supplemental table 3.**
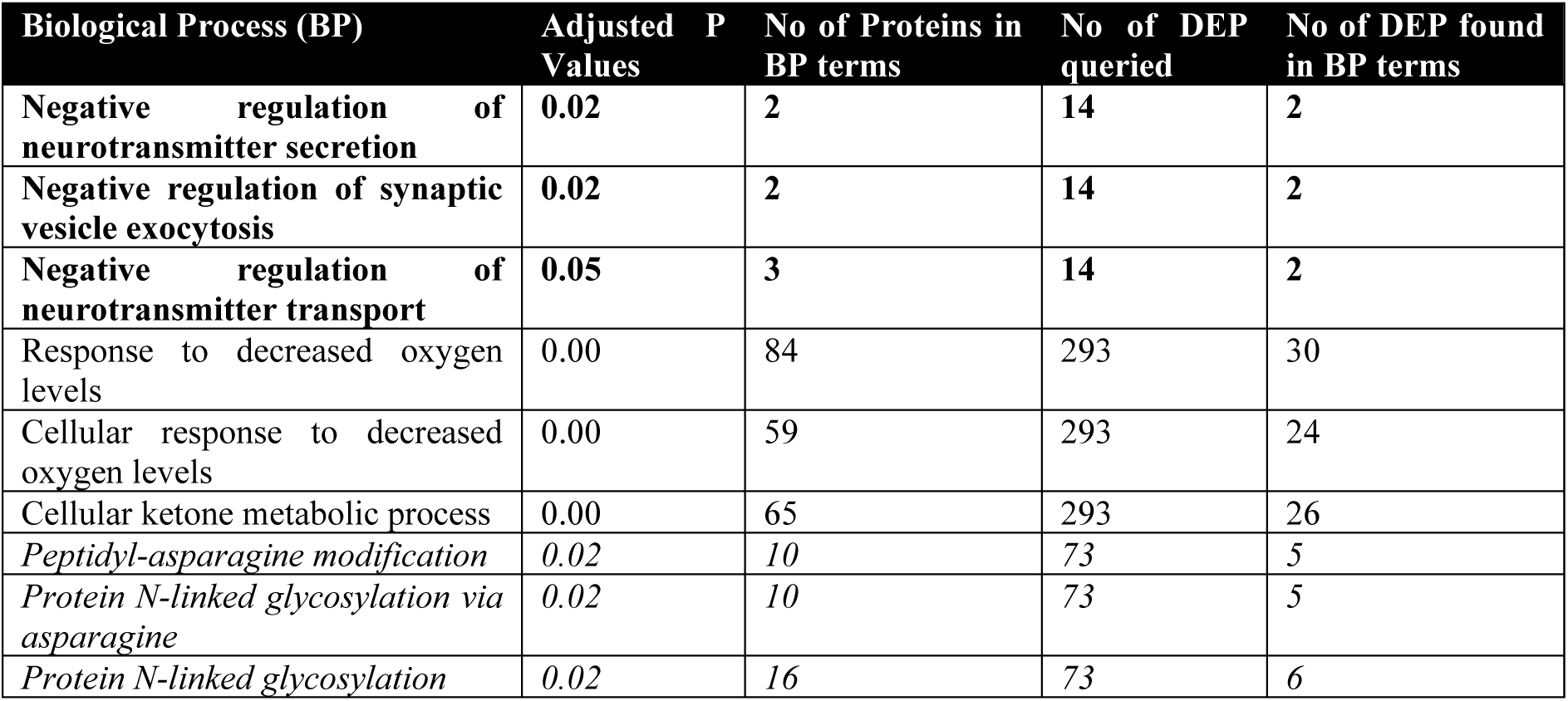
The top three functionally enriched gene ontologies are: biological process (GO: BP) Placenta (bold font), medium/large STB-EVs (normal font), and small STB-EVs (italics).

| Biological Process (BP) | Adjusted P Values | No of Proteins in BP terms | No of DEP queried | No of DEP found in BP terms |
| --- | --- | --- | --- | --- |
| **Negative regulation of neurotransmitter secretion** | **0.02** | **2** | **14** | **2** |
| **Negative regulation of synaptic vesicle exocytosis** | **0.02** | **2** | **14** | **2** |
| **Negative regulation of neurotransmitter transport** | **0.05** | **3** | **14** | **2** |
| Response to decreased oxygen levels | 0.00 | 84 | 293 | 30 |
| Cellular response to decreased oxygen levels | 0.00 | 59 | 293 | 24 |
| Cellular ketone metabolic process | 0.00 | 65 | 293 | 26 |
| *Peptidyl-asparagine modification* | *0.02* | *10* | *73* | *5* |
| *Protein N-linked glycosylation via asparagine* | *0.02* | *10* | *73* | *5* |
| *Protein N-linked glycosylation* | *0.02* | *16* | *73* | *6* |

**Supplemental table 4:**
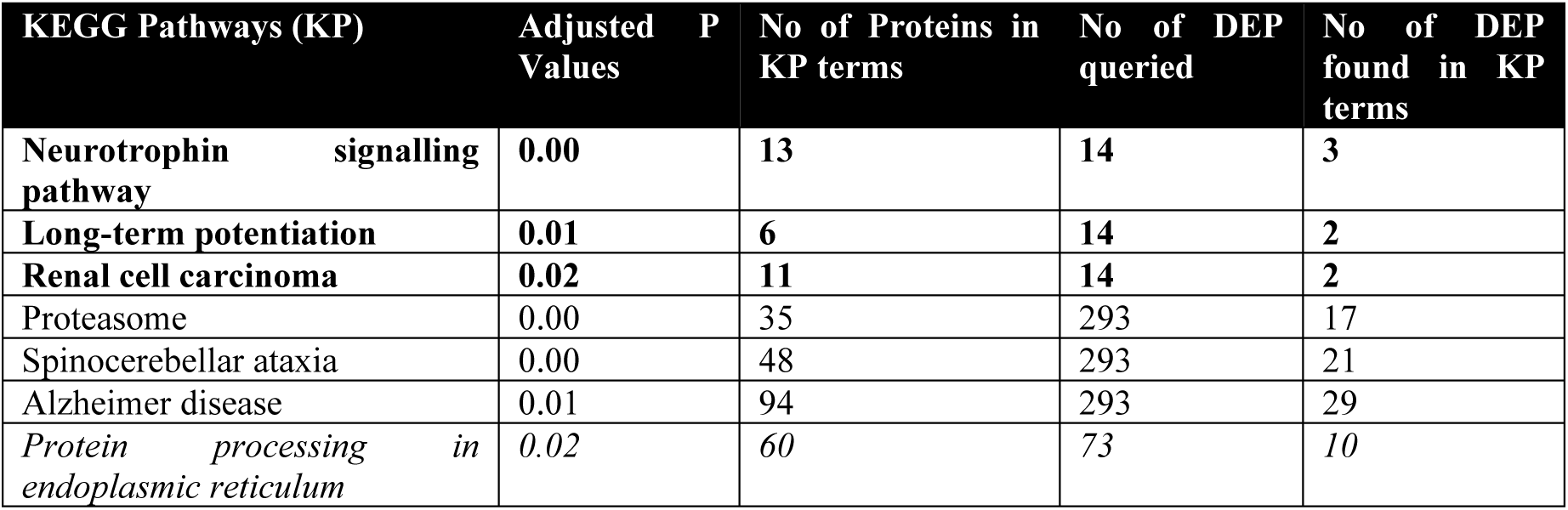
The top three functionally enriched KEGG Pathways in the placenta, medium/large STB-EVs, and small STB-EVs (only 1 KEGG). Placenta (bold font), medium/large STB-EVs (normal font), and small STB-EVs (italics)

### Major Resources Table

#### Antibodies

Major Resources Table Antibodies
| Target antigen | Vendor or Source | Catalog # | Working concentration | Dilution |
| --- | --- | --- | --- | --- |
| PLAP | In house antibody |  | 1.6 µg/µl | 1/1000 |
| CD63 | Santa Cruz Biotechnology | sc-365604 | 200 µg/µl | 1/1000 |
| ALIX | Cell Signalling | #2171 | 200 µg/µl | 1/1000 |
| Cytochrome C | Santa Cruz Biotechnology | sc-13560 | 200 µg/µl | 1/500 |
| SR-BI | Santa Cruz Biotechnology | ab52629 | 0.35 µg/µl | 1/1000 |
| Filamin B | Insight Biotechnology | GTX387 | 1.64 µg/µl | 1/1000 |
| PAPP-A2 | Abcam | ab59100 | 1 µg/µl | 1/1000 |
| Collagen 17A1 | Abcam | Ab28440 | 1 µg/µl | 1/2000 |
| Mouse Immunoglobulins | Dako UK Ltd | P044701 | 1 µg/µl | 1/2000 |
| Rabbit Immunoglobulins | Dako UK Ltd | P044801 | 1 µg/µl | 1/2000 |

#### Flow cytometry resources

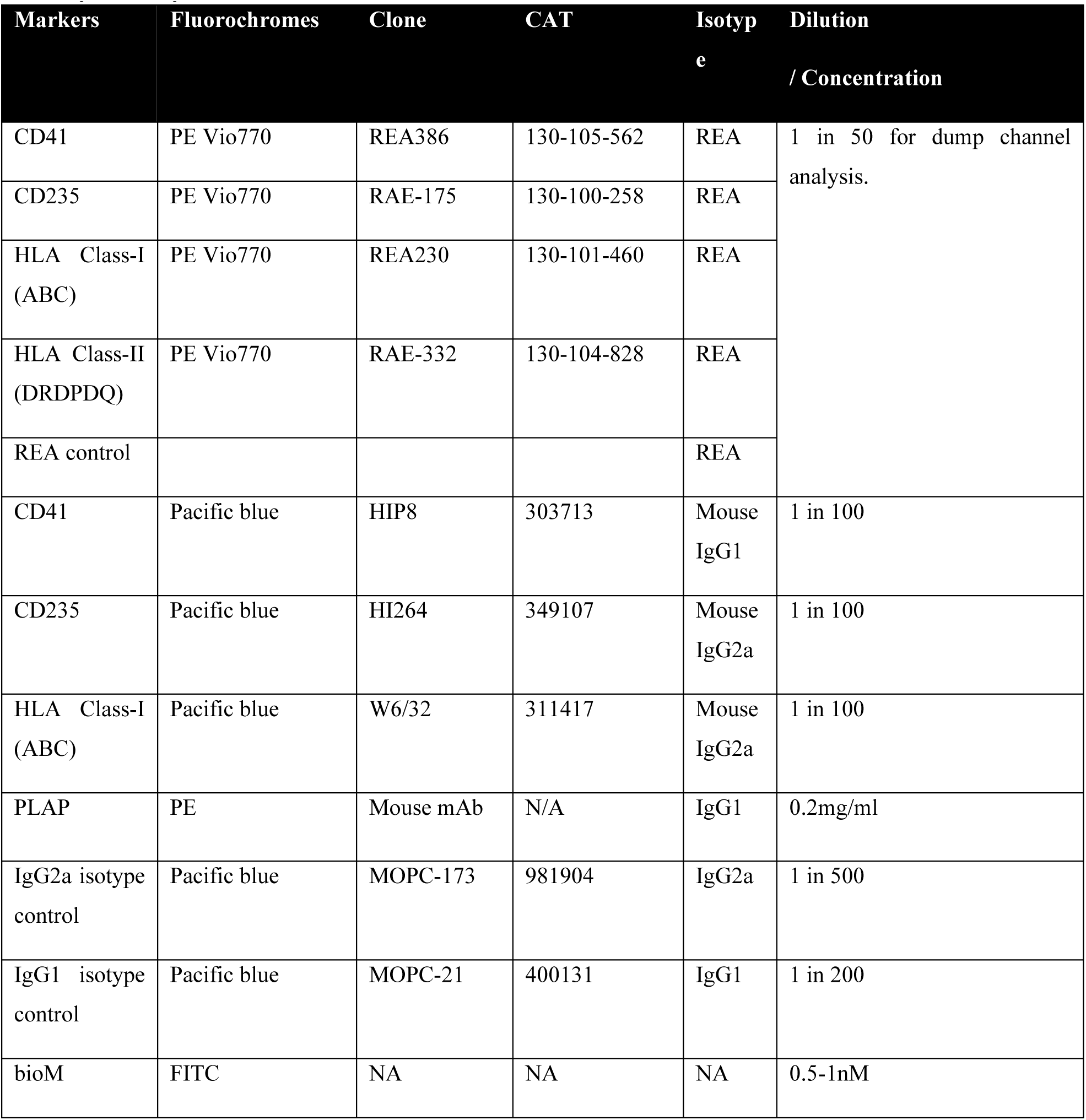

| Markers | Fluorochromes | Clone | CAT | Isotype | Dilution<br>/ Concentration |
| --- | --- | --- | --- | --- | --- |
| CD41 | PE Vio770 | REA386 | 130-105-562 | REA | 1 in 50 for dump channel analysis. |
| CD235 | PE Vio770 | RAE-175 | 130-100-258 | REA |  |
| HLA Class-I (ABC) | PE Vio770 | REA230 | 130-101-460 | REA |  |
| HLA Class-II (DRDPDQ) | PE Vio770 | RAE-332 | 130-104-828 | REA |  |
| REA control |  |  |  | REA |  |
| CD41 | Pacific blue | HIP8 | 303713 | Mouse IgG1 | 1 in 100 |
| CD235 | Pacific blue | HI264 | 349107 | Mouse IgG2a | 1 in 100 |
| HLA Class-I (ABC) | Pacific blue | W6/32 | 311417 | Mouse IgG2a | 1 in 100 |
| PLAP | PE | Mouse mAb | N/A | IgG1 | 0.2mg/ml |
| IgG2a isotype control | Pacific blue | MOPC-173 | 981904 | IgG2a | 1 in 500 |
| IgG1 isotype control | Pacific blue | MOPC-21 | 400131 | IgG1 | 1 in 200 |
| bioM | FITC | NA | NA | NA | 0.5-1nM |

